# HostSeq : A Canadian Whole Genome Sequencing and Clinical Data Resource

**DOI:** 10.1101/2022.05.06.22274627

**Authors:** S Yoo, E Garg, LT Elliott, RJ Hung, AR Halevy, JD Brooks, SB Bull, F Gagnon, CMT Greenwood, JF Lawless, AD Paterson, L Sun, MH Zawati, J Lerner-Ellis, RJS Abraham, I Birol, G Bourque, J-M Garant, C Gosselin, J Li, J Whitney, B Thiruvahindrapuram, J-A Herbrick, M Lorenti, MS Reuter, NO Adeoye, S Liu, U Allen, FP Bernier, CM Biggs, AM Cheung, J Cowan, M Herridge, DM Maslove, BP Modi, V Mooser, SK Morris, M Ostrowski, RS Parekh, G Pfeffer, O Suchowersky, J Taher, J Upton, RL Warren, RSM Yeung, N Aziz, SE Turvey, BM Knoppers, M Lathrop, SJM Jones, SW Scherer, LJ Strug

## Abstract

HostSeq was launched in April 2020 as a national initiative to integrate whole genome sequencing data from 10,000 Canadians infected with SARS-CoV-2 with clinical information related to their disease experience. The mandate of HostSeq is to support the Canadian and international research communities in their efforts to understand the risk factors for disease and associated health outcomes and support the development of interventions such as vaccines and therapeutics. HostSeq is a collaboration among 13 independent epidemiological studies of SARS-CoV-2 across five provinces in Canada. Aggregated data collected by HostSeq are made available to the public through two data portals: a phenotype portal showing summaries of major variables and their distributions, and a variant search portal enabling queries in a genomic region. Individual-level data is available to the global research community for health research through a Data Access Agreement and Data Access Compliance Office approval. Here we provide an overview of the collective project design along with summary level information for HostSeq. We highlight several statistical considerations for researchers using the HostSeq platform regarding data aggregation, sampling mechanism, covariate adjustment, and X chromosome analysis. In addition to serving as a rich data source, the diversity of study designs, sample sizes, and research objectives among the participating studies provides unique opportunities for the research community.

## BACKGROUND

Following exposure to SARS-CoV-2 (the virus that causes COVID-19), some individuals remain disease- or symptom-free while others develop a spectrum of symptoms from mild to severe with the potential for fatal outcomes (1). This variability in response to exposure suggests that susceptibility is mediated at least in part by host genetic factors (2). Genetic factors have been associated with acquisition and severity of other viral infections (3–7), including SARS-CoV-1 (8,9). A growing body of work demonstrates a role for host genetics in SARS-CoV-2 (10–14). Despite the relative novelty of the SARS-CoV-2 virus and the challenges of identifying genetic contributors in a changing environment (2), several loci contributing to infection susceptibility and illness severity have been identified (15). Associated loci are comprised of rare and common variations and occur throughout the genome, including but not limited to chromosome X and the HLA region on chromosome 6.

In 2020, several countries launched efforts to identify the genetic factors affecting COVID-19 outcomes to support diagnostics, therapy and vaccine development. However, Canada was not poised to do so because, although population-based cohorts exist (16,17), a national whole genome sequencing cohort broadly consented for research and translation, and linked to rich clinical and public health data, did not exist at the onset of the global pandemic. Here we describe the development of this national platform to address pressing questions concerning COVID-19 and other health outcomes in Canada. In April 2020, as part of the Canadian pandemic response, Genome Canada (a not-for-profit organization funded by the Government of Canada) launched the Canadian COVID-19 Genomics Network (CanCOGeN) (18). CanCOGeN established a coordinated pan-Canadian network of studies in collaboration with Canada’s national platform for genome sequencing and analysis (CGEn). Beginning June 2020, CGEn developed HostSeq: a national databank of independent clinical and epidemiological studies enrolling SARS-CoV-2-infected participants across Canada. The goal of HostSeq is to create a data repository with whole genome sequencing and harmonized clinical information, including comorbidities for 10,000 Canadians. With the launch of HostSeq, investigators can now begin to address questions of genetic susceptibility to SARS-CoV-2 infection and outcomes from the Canadian perspective. The approvals in place to link HostSeq to other local, provincial or national data resources expand the utility of the resource, including genetic susceptibility for future implications of SARS-CoV-2 infection. Further, summary statistics from association studies of HostSeq have been contributed and are aligned with international efforts including the COVID-19 Host Genetic Initiative (HGI) (19) and COVID Human Genetic Effort (https://www.covidhge.com/). Most importantly, we have established the research project infrastructure necessary for future pan-Canadian genome sequencing studies. In this resource paper introducing the HostSeq Databank, we present its design characteristics, high-level analytic considerations pertaining to it, and the research opportunities this rich resource provides.

## CONSTRUCTION AND CONTENT

### HostSeq Project Design

HostSeq (Figure 1) is a project representing a consortium of investigator-initiated SARS-CoV-2-related research studies across Canada. Each partner study was required to adhere to core consent elements (Table S1), contribute blood (or in rare cases saliva) samples for whole genome sequencing, and provide clinical information using a standardized case report form (Table S2).

**Figure 1.**
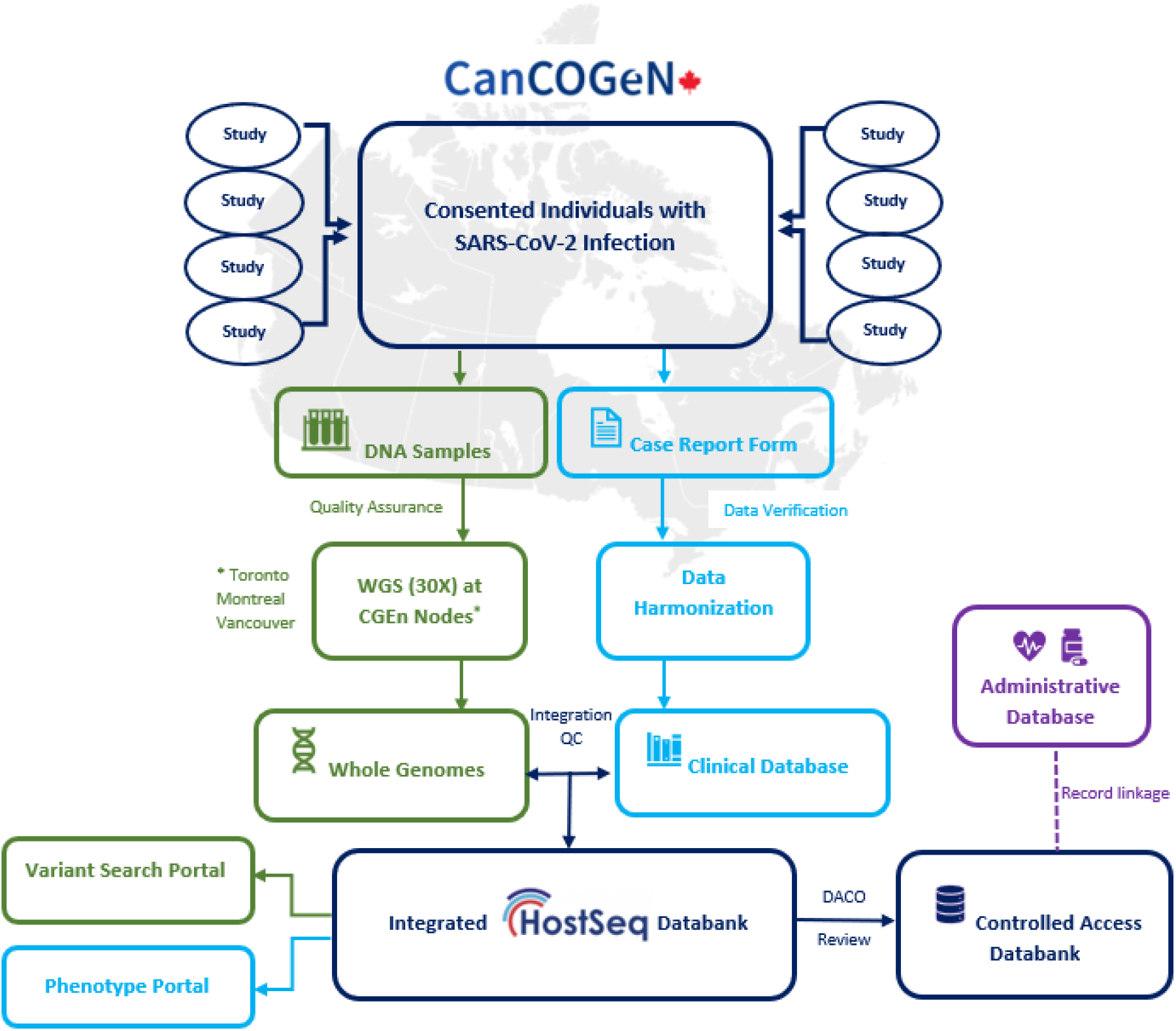
Sample and data flow in HostSeq ^1^.

Within these studies, eligible participants include individuals of any age with a positive SARS-CoV-2 test performed by any Health Canada approved method. In some studies, suspected cases with clinically assessed COVID-19-related symptoms but without a positive test diagnosis were also included. Within the primary studies, each participant consented to use of their whole genome sequence for future research (20). Participants also consented to the update, linkage and collection of their data from medical records and charts, as well as from administrative databases, and the deposition of data in a cloud-based, access-controlled databank which can be shared with approved researchers including international and commercial researchers. Additionally, participants had the option to consent to be re-contacted for updates or additional health information, or for invitations to participate in new research. Informed consent was obtained from individuals at each of the participating study sites. For the HostSeq Databank, approval was sought from the study’s Research Ethics Board (REB) for inclusion in HostSeq.

The HostSeq Databank shares data with the global research community following review and approval by the HostSeq-independent Data Access Compliance Office (DACO). The DACO verifies that the proposed research has REB approval from their host institution and conforms to HostSeq’s REB-approved SARS-CoV-2 or other health outcome research. DACO-approved researchers sign inter-institutional legal agreements, which outline how the shared data is to be used, stored, and privacy protected.

### Whole Genome Sequencing

All HostSeq samples undergo whole genome sequencing in a standardized fashion at one of the three CGEn nodes: Toronto (The Centre for Applied Genomics at The Hospital for Sick Children), Montréal (McGill Genome Centre at McGill University), and Vancouver (Canada’s Michael Smith Genome Sciences Centre) on the Illumina NovaSeq6000 platform at 30X depth. Prior to sequencing, quality assurance is performed at multiple stages throughout the process (21). Concordance of the genotyping pipeline among sequencing sites is verified using the Ashkenazi trio set from the Genome in a Bottle Consortium (22).

Sequenced samples are analyzed jointly using an in-house pipeline encoded in Nextflow (23) and Snakemake (31), containerized using Docker (21). The Genome Reference Consortium human build 38 (GRCh38 assembly version GCA_000001405.15) reference genome that includes the alternative HLA decoy genes^2^ is used. Genomes are processed following the Best Practices guidelines of the Genome Analysis ToolKit (GATK v4.2.5.0). This includes alignment of sequences to the reference genome, and the genotyping of each sample individually followed by joint-calling of all genotypes together. Associated scripts can be found in a public repository (https://svn.bcgsc.ca/bitbucket/users/jmgarant). Software packages used to process and analyze the WGS data are listed in Table S3.

The in-house pipeline is as follows. Sequences are aligned to the reference genome using DRAGEN mapper (DRAGMAP v1.3.0) (25), sorted with Picard tools (v2.25.0) and bases are recalibrated using the Base Quality Score Recalibration (BQSR) of GATK. GATK HaplotypeCaller is used in Dragen mode on diploid samples for short variant discovery. Aligned sequences are thus converted to genomic Variant Calling Format (gVCF) files, which are then filtered and imported to a GATK GenomicsDB for joint-calling using the GATK GenotypeGVCFs tool. We perform HLA Class I typing using OptiType software (v1.3.1) (26); perform housekeeping with bcftools (v1.11) and samtools (v1.14) (27); check for sample contamination using VerifyBamID2 (v2.0.1) (28); check agreement between reported sex-at-birth and sex chromosome composition using PLINK software (v1.90) (29); and predict ancestry admixture (30) and relatedness (31) using Genetic Relationship and Fingerprinting software (GRAF v2.4). We use PLINK (v2.00) (32) and R (3.6.3) (33) for genetic data analysis. Additionally, we compare the genetic principal components of HostSeq with the 1000 Genomes Project reference populations (34,35) following the guidelines of plinkQC (36). Samples are excluded based on the following checks (Figure S1): (i) genotyping call rate < 95%, (ii) sex chromosome composition and reported sex-at-birth mismatch, (iii) samples identified as duplicates, (iv) possibly mislabelled samples, (v) sample contamination rate > 3%, and (vi) mean coverage < 10. The whole genome sequence data are readily available in joint VCF format (aligned sequences can be made available upon request).

### Contributing Studies and Data Harmonization

As of December 20, 2022, 13 participating studies contributed data and biospecimens to HostSeq (Table S4). Although all 13 studies continue collecting clinical information, 6 have completed their participant recruitment. To date, we have harmonized data from all 13 studies. The participating studies are predominantly prospective SARS-CoV-2 studies based in hospitals, and are seeking to identify genetic factors that contribute to varying COVID-19 outcomes. Here we summarize characteristics of the 13 harmonized studies. Three studies—genMARK, Alberta Childhood COVID-19 Cohort (“AB3C”), and Genomic Determinants of COVID-19 (“GD-COVID”) —are using a case-control design, in which laboratory-confirmed COVID-19 cases are matched with controls (see Table S4 for matching factors and control eligibility). One study— Quebec COVID-19 Biobank (“BQC19”)—collected clinical data and biospecimens from 12 hospitals in Quebec (37). The remaining studies are case-cohorts with patients that either have a confirmed or suspected diagnosis of COVID-19. From these studies, the HostSeq Databank includes data from study subjects on demographics, comorbidities and assessment and treatment provided for COVID-19.

Clinical data from the participating studies is systematically harmonized by the HostSeq team in an ongoing process. In the first stage, we verify the raw data by checking for missingness, consistency, inadmissible values, and aberrant values across the variables. In the second stage, we harmonize the data guided by a set of common definitions and rules, including application of uniform classification, coding, and measurement units specified in the HostSeq Codebook (available through the *HostSeq Phenotype Portal* described below in *HostSeq Data Portals*). For example, all laboratory test variables are converted into predefined units; text entries in French are translated into English; and medications and complications variables are coded by timeline (prior to illness vs. during illness vs. post-discharge follow-up). Any potential data errors detected in the harmonization process are communicated to the participating study teams and resolved through follow-up.

Study-specific sample sizes currently range from 11 to 4,602. To date, in the HostSeq databank the 13 studies have contributed 9,913 clinical records and submitted 10,978 samples (Table 1). With the exception of two studies that have recruitment across multiple provinces (CANCOV, CONCOR-Donor; n=2,196), most studies are province-specific: six studies in Ontario (GENCOV, GenOMICC, SCB, LEFT-GEN, genMARK, Understanding Immunity to Coronaviruses; n=3,114), one in Quebec (BQC19; n=4,602), two in Alberta (AB3C; AB-HGS n=262) and two in British Columbia (GD-COVID, Host Factors; n=804). Table S4 summarizes their research objectives and study designs. Detailed information for each study is also provided on the CGEn website (https://www.cgen.ca/hostseq-studies-2).

**Table 1.** Status of DNA sample sequencing (as of December 20, 2022). SAMPLES column indicates DNA samples submitted to HostSeq for sequencing. DATA column indicates raw clinical records submitted to HostSeq. Of these, a total of 9,427 records have been harmonized.

| STUDY TITLE | STUDY |  |  |
| --- | --- | --- | --- |
|  | ACRONYM | SAMPLES | DATA |
| The Hospital for Sick Children's COVID-19 Biobank | SCB | 566 | 223 |
| Genetic Markers of Susceptibility to COVID-19 | genMARK | 876 | 738 |
| The Canadian COVID-19 Prospective Cohort Study | CANCOV | 1409 | 1,284 |
| The Genetics of Mortality in Critical Care | GenOMICC | 328 | 331 |
| Implementation of Serological and Molecular Tools to Inform COVID-19 Patient Management | GENCOV | 1,290 | 1,111 |
| Convalescent Plasma for COVID-19 Research | CONCOR-Donor | 787 | 787 |
| Host Genetic Factors Underlying Severe COVID-19 | Host Factors | 11 | 11 |
| The Quebec COVID-19 Biobank | BQC-19 | 4,602 | 4,323 |
| Genomic Determinants of COVID-19: Integration of Host and Viral Genomic Data to Understand the COVID-19 Epidemiologic Triangle | GD-COVID | 793 | 793 |
| HostSeq - Canadian COVID-19 Human Host Genome Sequencing Ottawa | LEFT-GEN | 43 | 43 |
| Understanding Immunity to Coronaviruses to Develop New Vaccines and Therapies against 2019-nCoV |  | 11 | 10 |
| Alberta Childhood COVID-19 Cohort Study | AB3C | 188 | 188 |
| Host Genetic Susceptibility to Severe Disease from COVID-19 Infection | AB-HGS | 74 | 71 |
| **TOTAL** |  | **10,978** | **9,913** |

## RESULTS

### Clinical Data Summary

The results discussed in this section are based on approximately 95% of the total expected cohort size of 10,000 participants. Although completeness varies across studies, we have achieved over 70% completeness of key variables capturing demographics, comorbidities, healthcare use, and patient outcome. Among the 9,427 currently available harmonized samples, HostSeq has 54.6% females and 41.5% males (and the remaining 3.9% are missing reported sex-at-birth), with an overall mean age (at recruitment) of 47.9 years. Distributions of sex and age vary across the studies (Table S5). Apart from studies including pediatric participants (AB3C, SCB), mean age in the studies ranges from 36.9 years (genMARK) to 63.5 years (GenOMICC). Underlying health conditions are collected in all studies, but using a variety of collection methods (medical chart reviews, participant surveys, and patient interviews). A total of 24 comorbidity variables across cardiovascular, respiratory, immunological, neurological systems, and other pathologies are collected in HostSeq. Distributions of comorbidities across the studies are available through the *HostSeq Phenotype Portal*.

While approximately half of the HostSeq participants were hospitalized and half were assessed in outpatient or community settings, the proportion of hospitalized versus non-hospitalized patients varied substantially across the studies. In all but one study (GenOMICC), participants presented predominantly with mild or moderate symptoms and did not require admission to intensive care units or invasive ventilation support. Of the hospitalized patients, 54.0% were discharged home, 15.0% were transferred to other hospitals or healthcare settings (e.g., rehabilitation centers or long-term care facilities) and 11.9% were reported deceased (Table 2).

**Table 2.** Hospitalization and patient outcomes in HostSeq. SD: standard deviation; ^1^ n=9,427 is a subset of the expected cohort of greater than 10,000; ^2^ Data not available for 605 participants (6.4%); ^3^ Data not available for 372 participants (3.9%); ^4^ Data not available for 595 participants (6.3%); ^5^Data not available for 1,721 participants (18.3%); ^6^ Denominator is 3,478 hospitalized participants. Data currently in collection for 652 participants (18.7%)

| HostSeq All Studies (n=9,427)<sup>1</sup> |  |  |
| --- | --- | --- |
| Age <sup>2</sup> | Mean (SD) | 47.9 (29.1) |
|  | Median (min, Q1, Q3, max) | 48.0 (0.1, 33.4, 61.9, 104.2) |
| Sex at birth <sup>3</sup> | Male | 3,911 (41.5%) |
|  | Female | 5,144 (54.6%) |
| Hospitalization <sup>4</sup> | Yes | 3,478 (36.9%) |
|  | No | 5,354 (56.8%) |
| ICU admission <sup>5</sup> | Yes | 1,148 (12.2%) |
|  | No | 6,558 (69.6%) |
| Patient outcome <sup>6,7</sup> | Discharged alive | 1,879 (54.0%) |
|  | Transfer to another facility | 521 (15.0%) |
|  | Palliative discharge | 3 (0.1%) |
|  | Hospitalized | 9 (0.3%) |
|  | Death | 414 (11.9%) |

### HostSeq Data Portals

HostSeq provides public access to two data portals: (1) The *Phenotype Portal* shows summaries for the major variables of the HostSeq harmonized clinical data; and (2) the *Variant Search Portal* enables queries in a genomic region to see all variants and their alleles identified in the HostSeq genomes. Both portals are static platforms that are updated periodically when a new release version of their respective data is available.

The *HostSeq Phenotype Portal* (https://hostseq.ca/phenotypes.html) provides information for clinical variables at aggregate and study-specific levels. Users can access variables by category (e.g., demographics, comorbidities, complications) and view their distributions (categorical variables are presented as boxplots, and numerical variables are presented as histograms and violin plots). Displays are limited to variables with ≥ 70% completeness. Researchers can also find links to the HostSeq study protocol and up-to-date data dictionaries on this portal.

The *HostSeq Variant Search Portal* (https://hostseq.ca/dashboard/variants-search) allows for queries of the HostSeq genetic data. The primary querying functionality is supported by the CanDIG-server (38), a platform enabling federated querying of genomics data. Beacon APIs (39) from the Global Alliance for Genomics and Health (GA4GH) are also built-in to allow HostSeq to join the federated Beacon network. Users can query information about a specific allele of interest. Information about the variants that can be queried includes their position and alleles and the respective internal frequencies of the alleles (minor allele frequencies are reported if they exceed 0.1). All columns in the table can be sorted and filtered.

### Genetic Data Summary

Results reported in this section are based on an interim joint-called set of 6,500 HostSeq genomes, of which 6,316 passed all quality checks (see *Methods*). Our predicted population structure covers five major ancestry groups (Figures 2 and S2-3; 69% European, 6% Admixed American, 8% East Asian, 8% South Asian, 6% African, and approximately 3% uncategorized) and closely matches self-reported ancestries (where available). Additionally, there are 300 and 518 pairs of first- and second-degree relationships, respectively.

**Figure 2.**
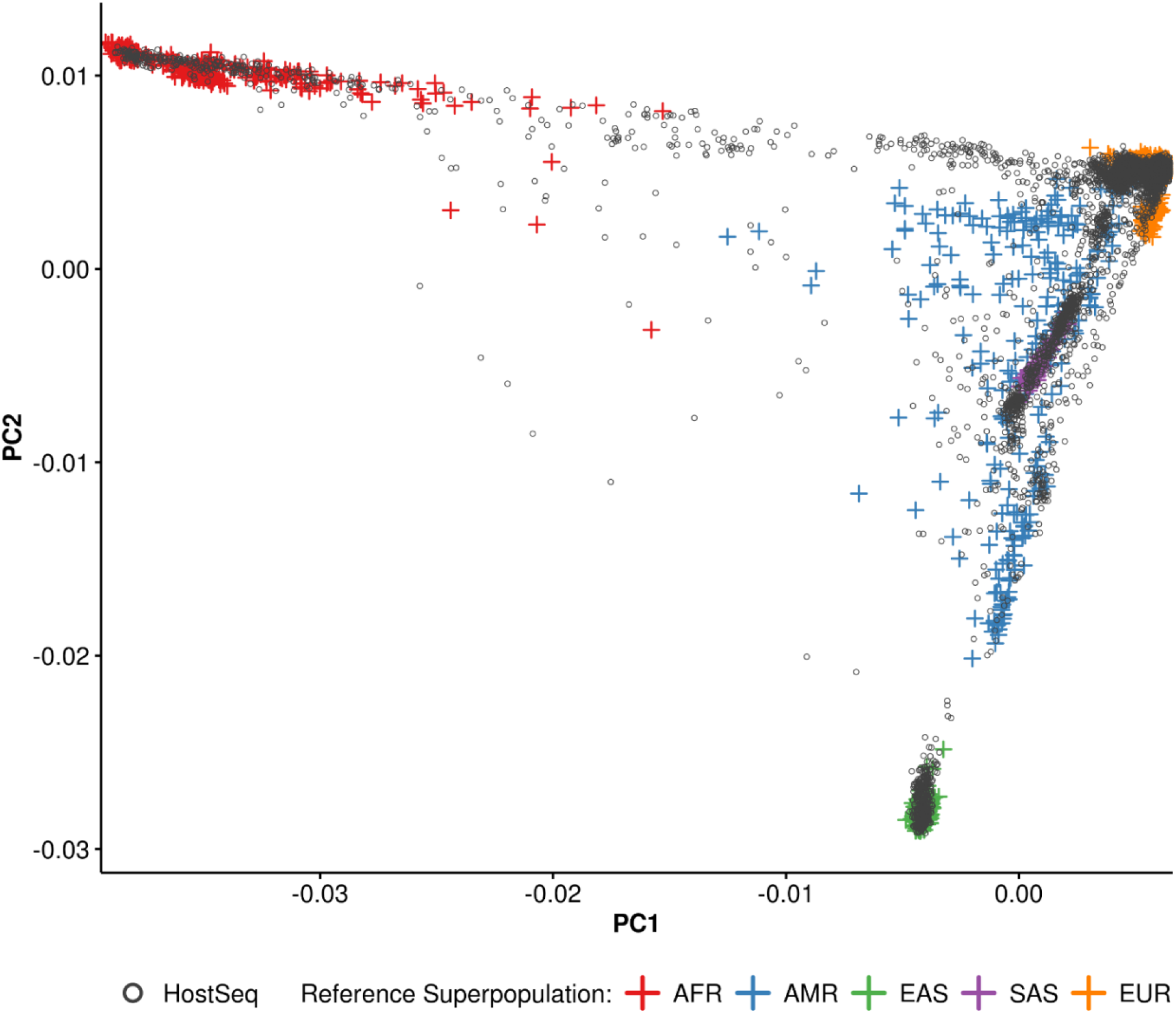
PCA projection of HostSeq genomes against reference superpopulations. HostSeq genomes were merged with the 1000 Genomes reference set. The first two principal components of this merged data are shown here with HostSeq genomes in black and 1000 Genomes samples colored by their superpopulation: AFR=African, AMR=Admixed American, EAS=East Asian, SAS=South Asian, EUR=European.

Currently HostSeq provides 174.5 million short variants consisting of single nucleotide variants and indels. We report HLA Class I haplotypes for three loci (*HLA-A, HLA-B* and *HLA-C*) with bi-allelic typing at 4-digit resolution (allele group with specific alleles). The numbers of unique alleles for *HLA-A, HLA-B* and *HLA-C* in 4436 genomes are 73, 145 and 49, respectively (the most common alleles per locus are *HLA-A\**02:01, *HLA-B\**07:02 and *HLA-C\**07:01).

## UTILITY AND DISCUSSION

HostSeq provides unique opportunities to explore the genetics among SARS-CoV-2 positive individuals in Canada and the facilitation of an organizational governance and oversight for researchers in Canada and beyond. Even though the participating studies in HostSeq are heterogenous with different designs and objectives (Table 3 and Table S4), HostSeq is an opportunity to leverage that diversity to address research questions. Several issues need to be considered when analysing HostSeq data in a given research context. For example: (1) whether data from different studies should be analysed separately or combined (and how to combine those data); (2) the selection strategies used by the contributing studies to recruit participants; (3) adjustment of covariates for association tests with genetic variants; and (4) the details of X chromosome analysis.

**Table 3.**
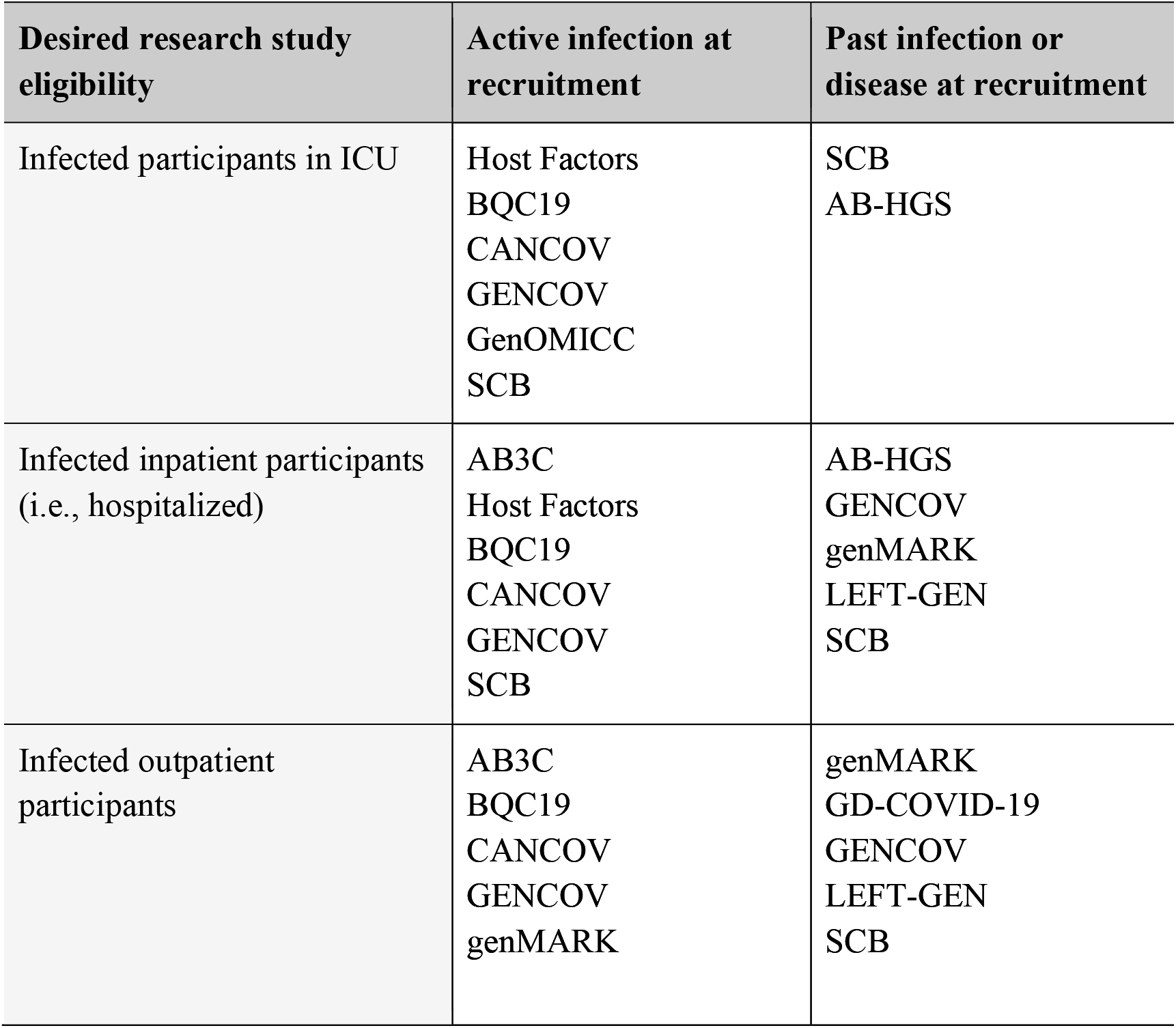
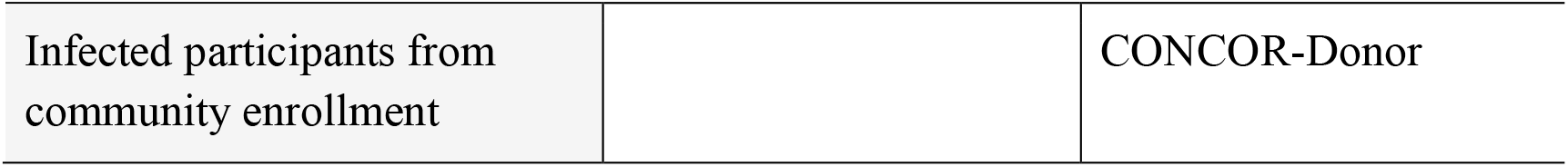
Aspects of participant ascertainment in HostSeq. Study participants who were not confirmed to be positive for infection either by molecular/serology test or clinical symptoms are not included in this Table.

### Individual or Combined Analysis

Whether an investigator’s research question would be best answered by within-study comparisons or analyses including multiple studies will require careful consideration of participant ascertainment criteria. For example, comorbidities might be analyzed within-study then combined via a meta-analysis to account for differences in study designs among the contributing studies. In contrast, for the disease severity indicated by hospitalization duration, it may be appropriate to jointly analyze the subset of studies that focus on in-patient recruitment. Table 3 provides details for the recruitment aspects that may frame such research questions. For example, to compare the genetics of hospitalized patients to non-hospitalized patients within the same study, data from AB3C, BQC19, CANCOV, GENCOV, genMARK, LEFT-GEN and SCB could be used. To compare ICU patients to non-ICU hospitalized patients, Host Factors, BQC19, CANCOV, GENCOV and SCB could be used.

Given the heterogeneity of the studies in HostSeq, the best approach for certain outcomes may be to analyse relevant studies individually. The feasibility of combining estimates or test results from separate studies, as in meta-analyses, depends on whether the individual studies measure and estimate the same features. The appropriateness of a joint analysis of participant data from multiple studies in an overarching model (perhaps with inclusion of study effects) also depends on whether the studies measure those same features. Although the combination of study-level estimates or tests can be as efficient as joint analysis in large samples (40), meta-analysis of summary data can be less efficient in smaller samples. When individual data are available, joint analysis is recommended, incorporating sparse-data methods for variants with low minor allele counts and outcomes with low prevalence (41,42). Furthermore, with study or environmental factors and other sources of heterogeneity, joint analysis can exploit gene-environment interaction (43) and give insight into sources of within- and between-study variation.

Given the dynamic nature of the COVID-19 pandemic, temporal and spatial variation within- and between-studies is another source of heterogeneity that is challenging and deserves consideration. Studies with prolonged recruitment and wide variation in dates of infection may allow such factors to be examined. When looking across the participating HostSeq studies, it may be of interest to examine changes in the profiles of recruited patients as the seropositivity rates and vaccination rates changed with time across Canada and as treatments changed and improved (for example, by combining HostSeq data with serological studies).

### Participant Selection Mechanism

Most of the participating studies are designed to include individuals who tested positive for SARS-CoV-2 at a participating institution or individuals who volunteered to donate blood and previously had a positive test. For such participants, it can be difficult to specify exactly what population they represent. To reduce bias and improve interpretation of results, the processes by which individuals join a given study needs to be considered (44). Here, we interpret bias relative to the effect of a variable (genetic or otherwise) in a target population. If an analysis is to involve an outcome variable (e.g., hospitalized versus not hospitalized), a genetic variable of interest and some additional covariates, then the validity of standard statistical methods is linked to how the sample inclusion depends on the outcome. Such dependence occurs in response-selective designs in which individuals are included in a study according to the values of an outcome (45–47). Except for the simple case-control setting, weighting or conditional estimation is needed to avoid estimation bias of the genetic association. Such methods require estimation or specification of the probability of being selected for inclusion. We encourage analyses that address study sample selection mechanisms.

Methods to account explicitly for selection conditions are similar to methods used for the analysis of secondary traits in case-control studies (48,49). From a methodological standpoint, we also encourage studies of bias and Type 1 error control when standard analyses are used (such as unweighted logistic regression). When the selection mechanism is not easily described, comparison of study samples to population or administrative data may provide insights.

Finally, as HostSeq includes various ancestries, care must be taken to avoid confounding through population stratification (for example, by use of stratification, mixed models, and genetic principal components). This issue, alongside issues related to the heterogeneity of participating studies, are not unique to HostSeq, and arise in most collaborative multi-center or consortium-based research.

### Covariate Adjustment

The choice of adjustment covariates in tests for association of outcome with a genetic variant is context dependent and open to discussion in many settings (50,51). In testing for genetic associations with COVID-19 outcomes, one strategy would be to adjust for factors such as age and sex that may affect selection or the outcome in question but are not associated with the genetic variant (unless it is on the sex chromosomes; as mentioned below in *Sex difference and X Chromosome Analyses* below). We must also consider whether to adjust for factors such as comorbidities, which may be related both to the outcome and to the variant. This is of particular importance for severe COVID-19: in the ICU, 1-year mortality outcomes increase with each additional week spent in ICU, each decade in age, and each additional comorbid illness in the Charlson score (52). From a causal perspective, adjusting for multiple covariates without a clear conceptual framework could lead to adjustment for variables that lie on the causal pathway (53). If there is a causal link from variant to outcome that passes through such a variable, then researchers could choose to test for either direct or indirect effects of the variant. As part of the process of learning about genetic effects on COVID-19 outcomes, we encourage analyses both with and without adjusting for such factors.

For the discovery stage in genetic association studies, power considerations are important. There have been suggestions that adjusting for too many covariates decreases power (51,54), and that two-phase strategies of genome-wide screening by simple analysis followed by targeted in-depth modelling is adequate and efficient. However, this is an area for which further study is warranted.

### Sex Difference and X Chromosome Analyses

COVID-19 displays sexual dimorphism with greater severity in males (55–57). In addition to environmental exposures and sex-specific autosomal genetic effects, it is reasonable to hypothesize that some X chromosomal variants play a role in COVID-19 outcomes. Indeed, one gene on the X-chromosome, the angiotensin-converting enzyme 2 (*ACE2*, Xp22.2), has been reported to be important in SARS-Cov-2 infection and genetic analysis has demonstrated association evidence with *ACE2* variants (19).

However, all published GWAS of SARS-CoV-2 susceptibility or COVID-19 severity, to the best of our knowledge, uses the traditional genotype coding (0, 1 and 2 for a female; 0 and 2 for a male) that assumes X-inactivation through a dosage compensation model (i.e., with alleles in the non-pseudo-autosomal regions being expressed exactly half of the time in genetic females (58)). Yet, it has been reported that close to one-third of the X chromosome genes can escape X-inactivation (59,60); if so, the genotype of a male should be coded 0 and 1 by convention. To robustly deal with X-inactivation uncertainty we recommend the use of recent methods for genetic analysis of SARS-CoV-2 related research questions such as model averaging and selection (61,62) and an easy-to-implement regression model (63). Rare X-chromosome variant analysis (64,65) and X-inclusive polygenic risk scores also require careful consideration and further research.

### Health Research in the Canadian Context

People living in Canada are insured under single-payer health care systems administered at the provincial or territorial level. These systems broadly cover physician and hospital services, as well as procedures. This provides a unique opportunity to conduct passive follow-up to understand the short-term and long-term outcomes related to SARS-CoV-2 infection. Administrative health data are generated through patient contact with the health care systems and maintained in multiple databases that, with the appropriate approvals, can be linked using a unique encoded identifier to study specific, patient-level data (including genetic data). These data are administrative or procedural (e.g., surgeries, emergency department visits, hospital visits, comorbidities, routine medical exams), clinical (e.g., prescription medications, cancer screening), laboratory (e.g., blood measurements), social (e.g., education, income), and environmental (e.g., rurality, walkability, food insecurity, exposure to air pollution). The participant informed consent used by HostSeq allows for linkage to these data, transforming the HostSeq dataset into a longitudinal study. Specifically, linkage to administrative provincial data will provide: 1) a retrospective, longitudinal account of medical histories, health system utilization and diagnoses; and 2) prospective, longitudinal follow-up tracking the natural history of SARS-CoV-2 infection including multisystem inflammatory syndrome in children (MIS-C) and Long COVID, identifying new diagnoses (e.g., diabetes, cancer), long-term health outcomes (e.g., premature mortality), and health resource utilization. Linkage of the HostSeq study samples to provincial administrative data offers opportunities to collect additional data on risk factors and longitudinal outcomes, and opportunities to extend genetic association analyses. Administrative data can also facilitate evaluation of the representativeness of study samples and inform future study design.

The limitations of HostSeq data for investigation of specific scientific questions depend on limitations of the relevant participant studies. In addition, investigations that involve combining data or results from separate participant studies may require assumptions about comparability or heterogeneity; such assumptions should be scrutinized.

## CONCLUSIONS

Through the HostSeq initiative, Canada has built research infrastructure to investigate health effects of SARS-CoV-2 infection and COVID-19, and their association with genetic variants. This infrastructure can also be used for future epidemics. The unique features of the HostSeq project highlighted here present novel opportunities to develop, evaluate, and apply statistical methods that contribute to the understanding of genetic associations with COVID-19-related morbidity and mortality, as well as other phenotypes. The augmentation and linkage of the HostSeq questionnaire and genetic databank with other data resources is made possible by broad and flexible consent and will generate a dynamic population-based resource. This will allow for study of a broad range of research questions and sustained productivity over the years to come.

## Data Availability

All data produced in the present study are available through a Data Access Agreement and Data Access Compliance Office approval.

http://www.cgen.ca/daco-main

https://www.cgen.ca/hostseq-studies-2

https://hostseq.ca/phenotypes.html

https://hostseq.ca/dashboard/variants-search

## DECLARATIONS

### Ethics approval and consent to participate

HostSeq was approved by the Research Ethics Board of the Hospital for Sick Children (lead site) (#1000070720 from 2020-present). Written informed consent was obtained from all participants or parents/guardians/substitute decision makers prior to inclusion in the study. Additional REB information from the participating PIs:

| Site | PI | REB |
| --- | --- | --- |
| The Hospital for Sick Children | Stephen Scherer | 1000070720 |
| The Hospital for Sick Children | Rae Yeung | 1000070060 |
| The Hospital for Sick Children | Upton Allen | 1000069580 |
| University Health Network | Angela Cheung | CTO ID 2157 |
| Queens University | David Maslove | CTO ID 3209 |
| Mount Sinai Hospital | Jordan Lerner-Ellis | CTO ID 3302 |
| The Hospital for Sick Children | Rulan Parekh | 1000070462 |
| BC Children's Hospital | Catherine Biggs | H20-01667 |
| University of Montreal Health Centre |  | 19.389 (internal) MP-02-2020-8929 (multicentre) |
| BC Children's Hospital | Stuart Turvey | H21-00054 |
| The Ottawa Hospital | Juthaporn Cowan | 20210119-01H |
| Unity Health | Mario Ostrowski | 20-044 |
| University of Calgary | Gerald Pfeffer | 20-0574_MOD3 |
| University of Calgary | Francois Bernier | 20-0480_MOD6 |

### Consent for publication

Not applicable.

### Availability of data and materials

The datasets generated and/or analysed during the current study are not publicly available due to privacy concerns but are available through a Data Access Agreement and Data Access Compliance Office (DACO) approval (https://www.cgen.ca/daco-main). Aggregated data are publicly available through two data portals: a phenotype portal showing summaries of major variables (https://hostseq.ca/phenotypes.html) and their distributions, and a variant search portal enabling queries in a genomic region (https://hostseq.ca/dashboard/variants-search). The code used for processing the WGS data can be found in a publicly accessible repository (https://svn.bcgsc.ca/bitbucket/users/jmgarant).

### Funding

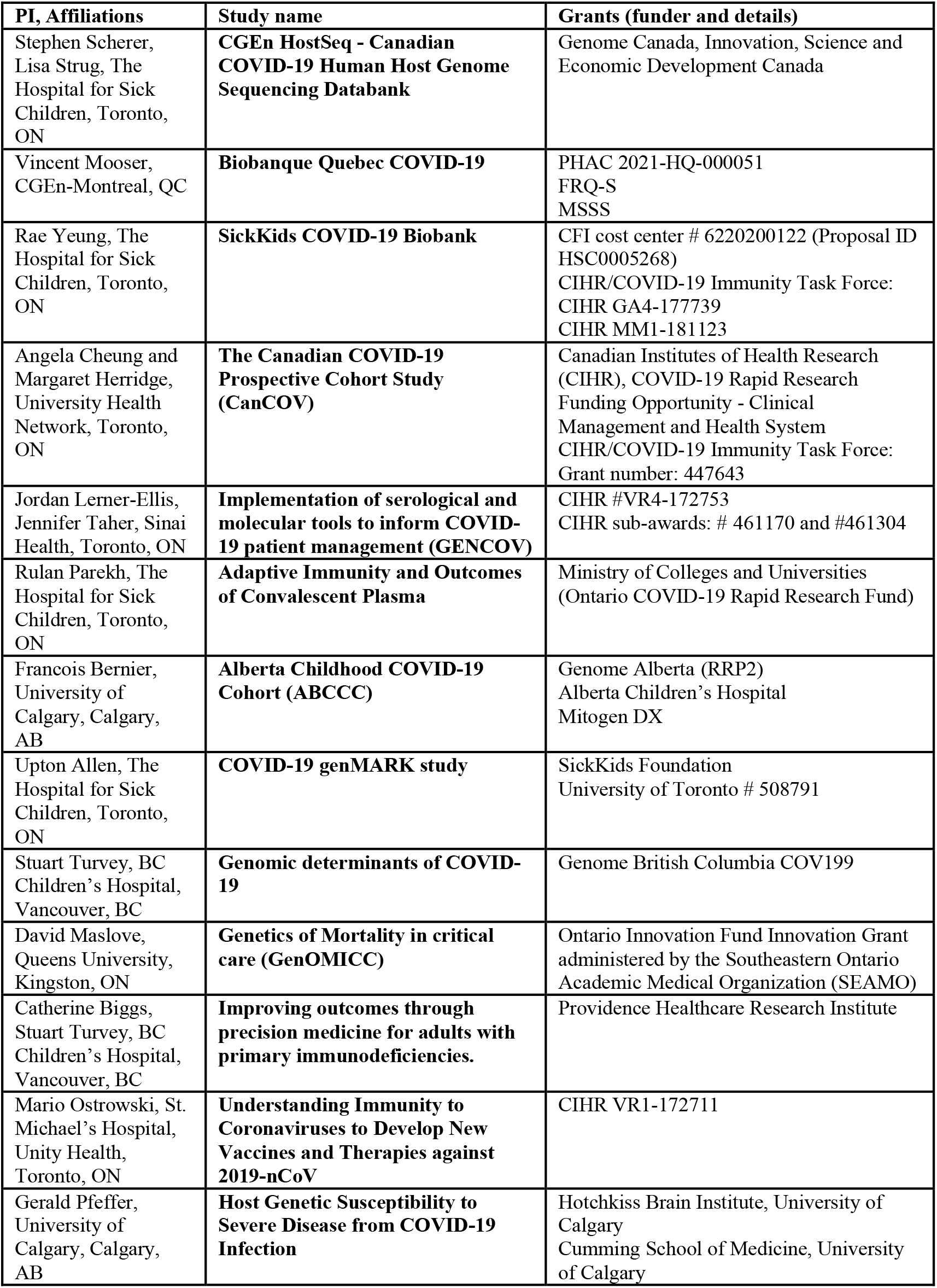

### Authors’ contributions

LJS led the study design and implementation.

Data harmonization: SY; genomic data analysis: EG.

Genetic epidemiology: LJS, JDB, SBB, LTE, FG, CMTG, RJH, JFL, ADP, LS.

Writing: SY, EG, LTE, RJH, ARH, JDB, SBB, FG, CMTG, JFL, ADP, LS, LJS.

Data processing and sharing: SY, EG, LTE, MLo, ARH, RJSA, IB, GB, J-MG, CG, JL, JW, BT, MSR, J-AH, NOA, SL, MHZ; SJMJ led the data processing and sharing.

Study contributors: JL-E, UA, FPB, CMB, AMC, JC, MH, DMM, BPM, VM, SKM, MO, RSP, GP, OS, JT, SET, JU, RLW, RSMY.

NA coordinated the three CGEn nodes; SET led study site recruitment; BMK designed the consent and data access process; MLa led the Quebec site; SJ led sequence informatics and the variant portal; NA, SWS and LJS provided overall study oversight.

## Acknowledgements

We wish to express gratitude to all HostSeq project participant studies and the individual participants within these studies for their contribution.

## APPENDIX

**Table S1.** HostSeq Core Consent Elements. In order to deposit datasets in <u>HostSeq COVID-19 controlled-access Databank,</u> all the elements in this table must be obtained in the research consen**t**.

| ELEMENTS | DETAILS |
| --- | --- |
| **Research data** | Whole genome sequencing of the sample and the ongoing collection of clinical data from participant's medical records/chart, administrative databases, etc. |
| **International sharing** | International sharing of genetic and clinical data |
| **Future research use** | Future health research on COVID-19 and other health outcomes |
| **Commercial use** | Use of genetic and clinical data for commercial purposes |
| **Controlled access** | Sharing of genetic and clinical data through a controlled-access mechanism |
| **Storage on cloud servers in Canada** | Storage of genetic and clinical data in the HostSeq Databank, on centralized Canadian cloud servers |
| **Duration of storage** | Indefinite storage of genetic and clinical data |
| **Data withdrawal** | Not possible to withdraw data that has already been distributed and used |
| **Re-identification** | Low risk that the participant could be re-identified in the future |
| **Re-contact (optional)** | Optional re-contact to update personal information, provide new health information, or to be invited to participate in new research projects. |

**Table S2.** HostSeq Case Report Form.

| IDENTIFICATION | COMORBIDITIES | LAB RESULTS |
| --- | --- | --- |
| Host Hospital | **Comorbidities: Immune system** | Haemoglobin |
| Source study participant ID | HIV | WBC count |
| HostSeq ID | Immunocompromised status | Lymphocyte count |
| DNA ID | Onset of COVID-19 to immunocompromisation | Neutrophil count |
| Sequencing site | Organ transplant | Haematocrit |
| Was informed consent obtained? | Organ type of transplant | Platelets |
| Study enrollment date | Autoimmune or rheumatologic disease | APTT/APTR |
| DOB | Diabetes | APTT |
| Age | Type I diabetes | PT |
| Laboratory COVID-19 test result | Type II diabetes | INR |
| Suspected to be COVID-19 positive? | **Comorbidities: Respiratory system** | ALT/SGPT |
| Ambulatory | Asthma | Total Bilirubin |
| Hospitalized State | COPD | AST/SGOT |
| Date of admission | Cystic fibrosis | Glucose |
| Sex at birth | Sleep apnea | Blood Urea Nitrogen (urea) |
| Gender | Do you use a home CPAP device? | Lactate |
| Ancestry | **Comorbidities: Genitourinary/Metabolic** | Creatinine |
| Height (cm) | Chronic kidney disease | Sodium |
| Weight (Kg) | Liver disease | Potassium |
| **DEMOGRAPHICS** | Gallbladder | Procalcitonin |
| Country of birth | Pancreas | CRP |
| Education | **Comorbidities: Cardiovascular system** | CT chest |
| Employment | Coronary intervention | Chest X-Ray performed? |
| Type of residence | Coronary artery bypass | ECG |
| Household composition | Congestive heart failure | POCUS |
| **PREGNANCY** | Hypertension | Echocardiogram |
| Pregnancy - currently pregnant? | Myocardial infarction | LDH |
| **RISK FACTORS** | Myocardial infarction Type I | D-Dimer |
| Cigarettes | Myocardial infarction Type II | Fibrinogen |
| How many cigarettes daily? | Peripheral vascular disease | Ferritin |
| Vaping | Stroke | Triglycerides |
| Cannabis | Arrhythmias | IL-6 |
| **AT ADMISSION/ASSESSMENT** | **Comorbidities: Neurological** | CD4 |
| COVID-19 test date | Dementia | CD8 |
| Date of diagnosis | Neurological or neuropsychiatric disease | CD4/CD8 ratio |
| Serology | **Comorbidities: Cancer** | NT-proBNP |
| Serology positive test date | Currently diagnosed with cancer? | BNP |
| Date of last negative test | Patient age at diagnosis | Troponin |
| PCR | Leukemia | **TREATMENT** |
| Commercial serology test kit name | Lymphoma | ICU or High Dependency Unit admission? |
| Onset date of first/earliest symptom | Sarcoma | If yes, date of ICU admission |
| ER triage date at this facility | Carcinoma | If yes, date of ICU discharge |
| Transfer from another facility? | Myeloma | Prone ventilation |
| ABO blood type | Mixed types | Inhaled nitric oxide |
| Rh factor | Cancer location | Tracheostomy inserted |
| Home medications | Cancer treatment in the past 12 months | Extracorporeal support |
| BCG vaccine | Other comorbidities | RRT or dialysis |
| COVID-19 Vaccine? | **PATHOGEN TESTING** | Inotropes/vasopressors |
| Name of first dose vaccine | Pathogen testing done during this illness | OTHER intervention or procedure |
| Name of second dose vaccine | Influenza | **MEDICATION** |
| **SYMPTOMS** | Coronavirus | Antiviral agent |
| Cough | RSV | Azithromycin “Zithromax” |
| Days with cough | Adenovirus | Any other antibiotic |
| Difficulty breathing | Enterovirus | Corticosteroid |
| Fever | Bacteria | Antifungal agent |
| Days with fever | Other infectious respiratory diagnosis | Colchicine |
| Heart rate | Was there a physician diagnosis of pneumonia? | Chloroquine “Aralen” |
| Highest respiratory rate | Suspected non-infective | Hydroxychloroquine |
| Systolic blood pressure | **COMPLICATIONS** | Tocilizumab “Actemra” |
| Diastolic blood pressure | Viral pneumonitis | Kineret “Anakinra” |
| Oxygen saturation | Bacterial pneumonia | IVIG |
| Fatigue | Acute Respiratory Distress Syndrome | Plasma |
| Myalgia (general aches and pain) | Pneumothorax | Other COVID therapy |
| Runny nose | Pleural effusion | **OUTCOME** |
| Sore throat | Cryptogenic organizing pneumonia (COP) | Outcome |
| Loss of taste/smell sense | Bronchiolitis | Outcome date |
| Nosebleed | Meningitis / Encephalitis | Self-care ability versus before illness |
| Ear pain | Seizure | Repeat hospital visit within 30 days? |
| Wheezing | Stroke / Cerebrovascular accident | Date and reason |
| Chest pain | Congestive heart failure |  |
| Joint pain | Cardiac inflammation (mark all that apply) |  |
| Headache | Cardiac arrhythmia |  |
| seizures | Cardiac ischaemia |  |
| Altered consciousness/confusion | Cardiac arrest |  |
| Abdominal pain | Coagulation disorder / Disseminated Intravascular Coagulation |  |
| Diarrhea | Anemia |  |
| Nausea/vomiting | Rhabdomyolysis / Myositis |  |
| Conjunctivitis | Acute renal injury/ Acute renal failure |  |
| Skin rash | Gastrointestinal haemorrhage |  |
| Other symptoms | Pancreatitis |  |
| Asymptomatic | Liver dysfunction |  |
|  | Hyperglycemia |  |
|  | Hypoglycemia |  |
|  | Inflammatory syndrome/Kawasaki Disease like |  |
|  | Other, please specify |  |

**Table S3.** Software used for processing WGS data.

| PROCESS | SOFTWARE | VERSION | URL |
| --- | --- | --- | --- |
| Sequence alignment | DRAGMAP | 1.3.0 | https://github.com/Illumina/DRAGMAP |
| Sorting alignments | Picard tools | 2.25.0 | https://broadinstitute.github.io/picard/ |
| Genotyping | HaplotypeCaller | GATK 4.2.5.0 | https://gatk.broadinstitute.org/hc/en-us/articles/4418062719899-HaplotypeCaller |
| Joint-calling | GenotypeGVCFs | GATK 4.2.5.0 | https://gatk.broadinstitute.org/hc/en-us/articles/4418054384027-GenotypeGVCFs |
| HLA Class I typing | OptiType | 1.3.1 | https://github.com/FRED-2/OptiType |
| Sample contamination | VerifyBamID2 | 2.0.1 | https://github.com/Griffan/VerifyBamID |
| Ancestry and sex prediction | GRAF | 2.4 | https://github.com/ncbi/graf |
| Miscellaneous | GATK | 4.2.5.0 | https://gatk.broadinstitute.org/hc/en-us/articles/4418051394587--Tool-Documentation-Index |
|  | Samtools | 1.14 | http://samtools.github.io/ |
|  | Bcftools | 1.11 | http://samtools.github.io/bcftools/bcftools.html |
|  | PLINK | 1.90 and 2.0.0 | https://www.cog-genomics.org/plink/ |
|  | R | 3.6.3 | https://cran.r-project.org/ |

**Table S4.**
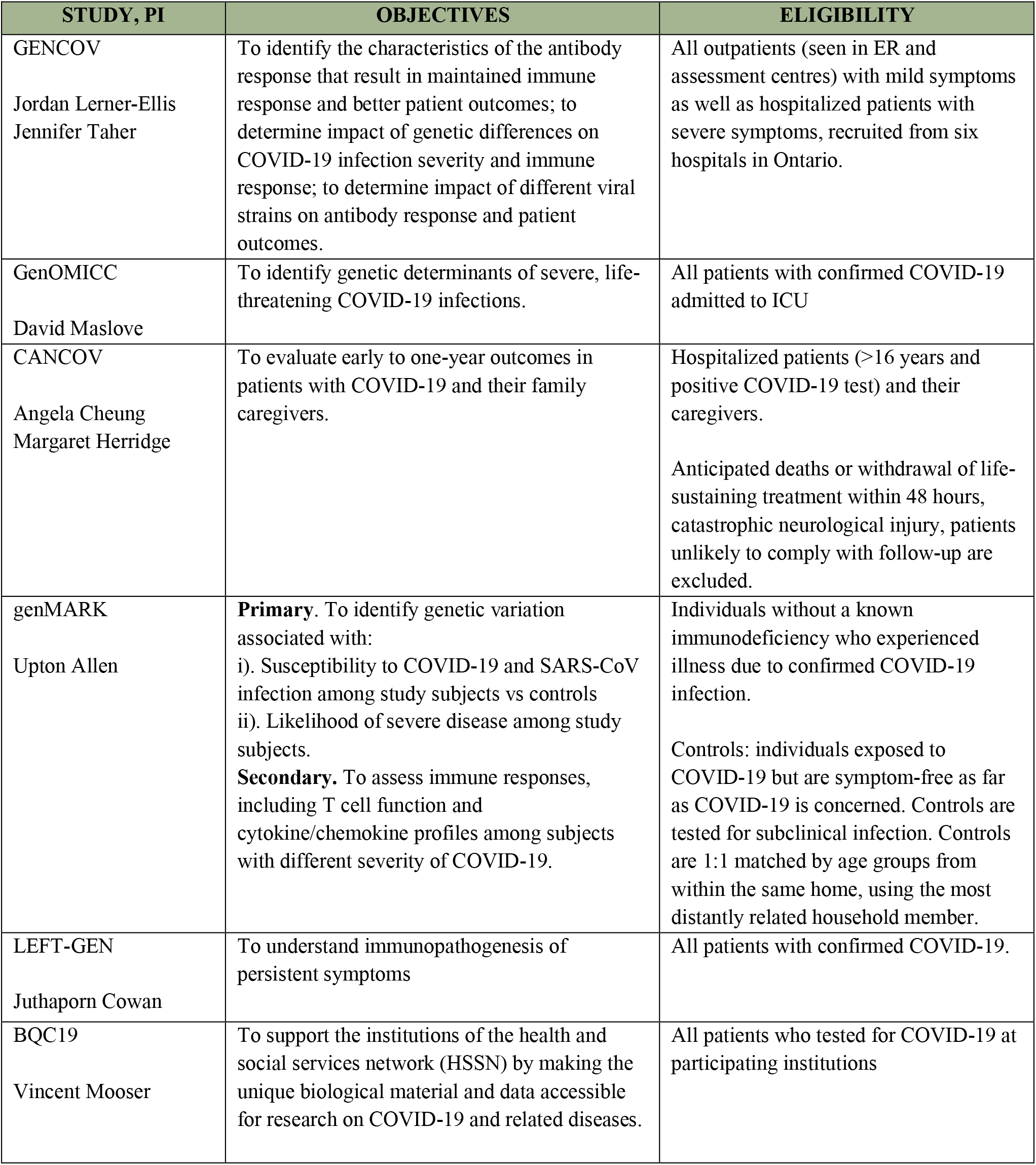
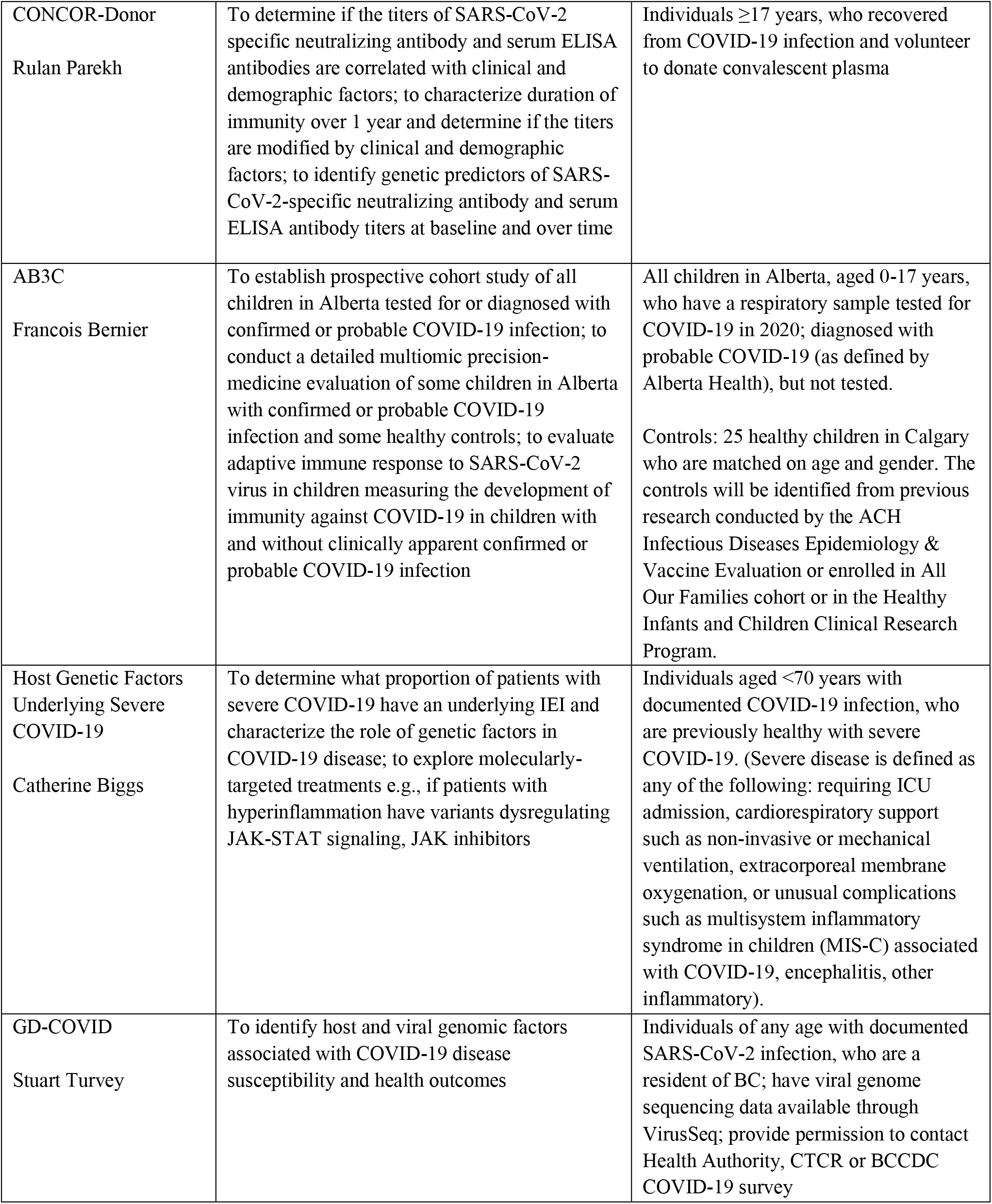
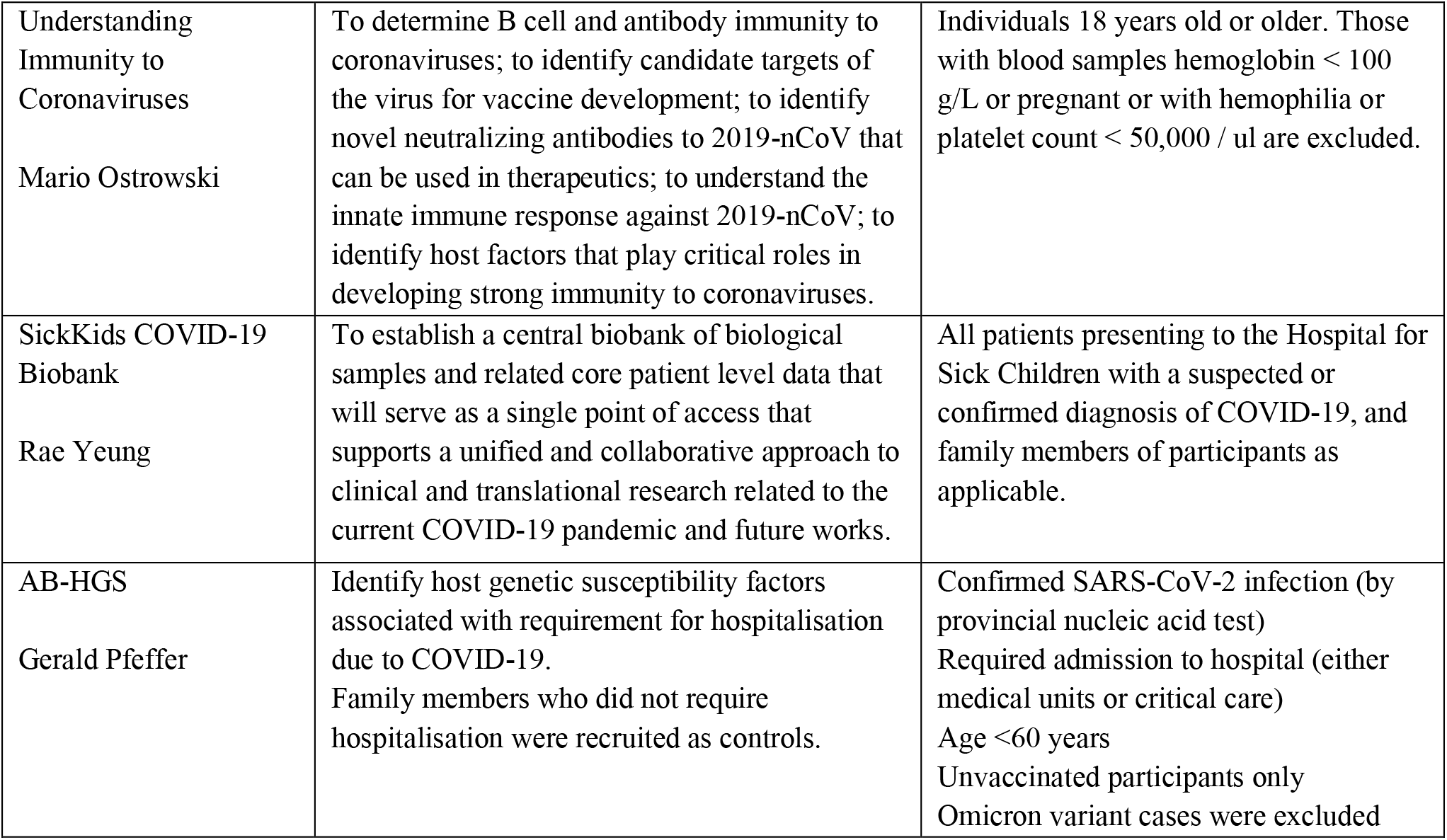
List of HostSeq participating studies as described in respective protocols.

**Table S5.** Distribution of sex and age across HostSeq studies (n=9,427). SD: Standard deviation; IQR: interquartile range.

| STUDY | SEX |  | AGE |  |
| --- | --- | --- | --- | --- |
|  | Male | Female | Mean (SD) | Median (IQR) |
| GENCOV | 515 (46.4%) | 596 (53.6%) | 45.3 (14.6) | 44.0 (23.0) |
| GenOMICC | 179 (54.1%) | 152 (45.9%) | 63.5 (14.4) | 66.0 (18.5) |
| CANCOV | 558 (42.8%) | 746 (57.2%) | 51.8 (16.1) | 51.0 (24.0) |
| CONCOR-Donor | 241 (30.2%) | 557 (69.8%) | 44.5 (13.5) | 44.0 (22.0) |
| BQC19 | 1,744 (46.4%) | 2,011 (53.6%) | 52.5 (20.6) | 52.5 (29.5) |
| genMARK | 170 (28.5%) | 426 (71.5%) | 36.9 (15.8) | 38.0 (21.0) |
| LEFT-GEN | 23 (52.3%) | 21 (47.7%) | 50.9 (15.5) | 50.0 (24.3) |
| GD-COVID-19 | 308 (38.8%) | 485 (61.2%) | 46.3 (18.9) | 46.0 (28.0) |
| Host Genetic Susceptibility | 36 (50.7%) | 35 (49.3%) | 49.5 (11.4) | 52.0 (16.0) |
| Understanding immunity | 5 (50.0%) | 5 (50.0%) | 43.6 (13.9) | 42.5 (21.0) |
| AB3C | 95 (50.5%) | 93 (49.5%) | 11.5 (4.5) | 12.0 (7.0) |
| SCB | 144 (64.6%) | 79 (35.4%) | 9.6 (7.0) | 8.0 (13.0) |

**Figure S1.**
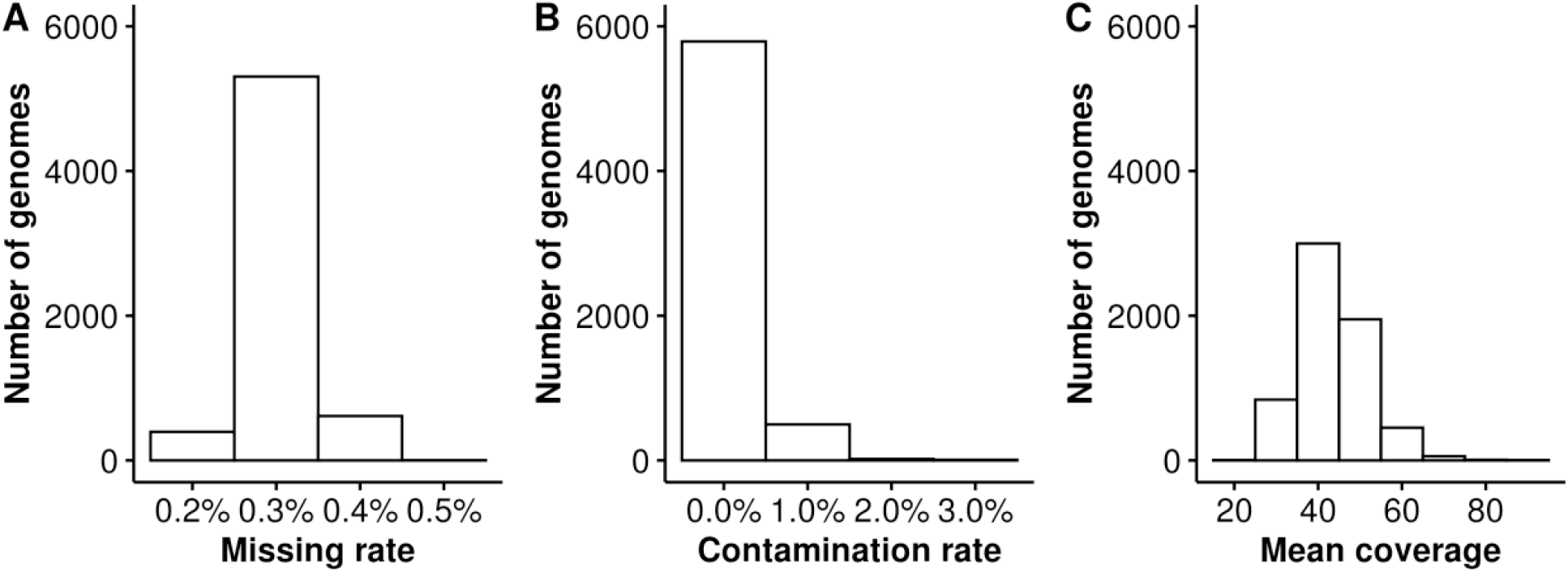
Quality of HostSeq genomes. (A) Missing rate < 5% (B) Contamination rate < 3% (C) Mean coverage >10.

**Figure S2.**
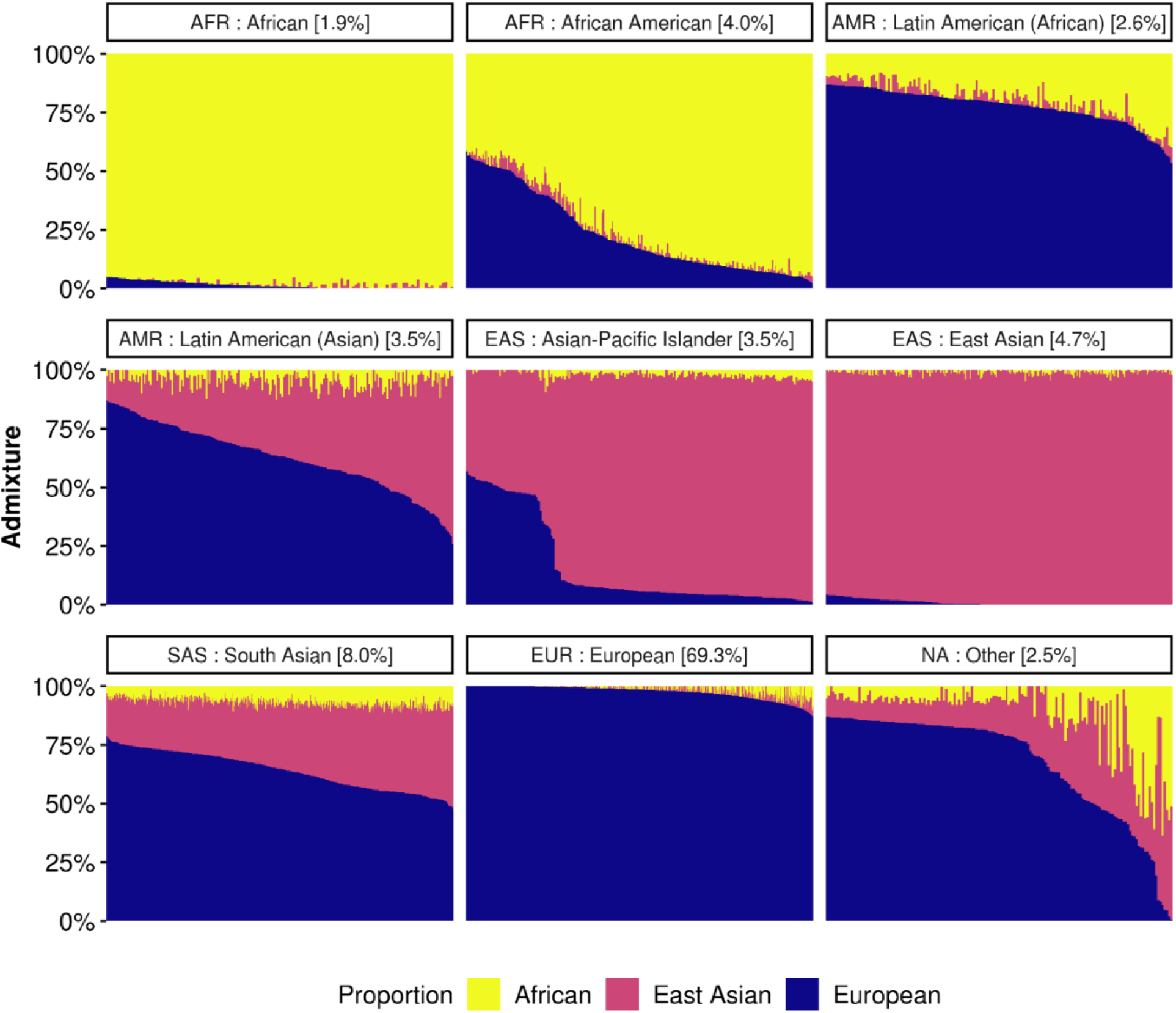
Predicted population admixture and ancestry classification in HostSeq genomes. Each bar represents a genome. Proportion of African, East Asian and European ancestries is determined (according to their definitions in GRAF), and genomes classified into 8 ancestry groups using GRAF-pop (see Methods). They are further categorized into 5 superpopulations: (i) AFR - African and African-American, (ii) AMR - Latin American Asian and Latin American African, (iii) EAS - Asian-Pacific Islander and East Asian, (iv) SAS - South Asian, and (v) EUR - European. 3% of genomes remain uncategorized.

**Figure S3.**
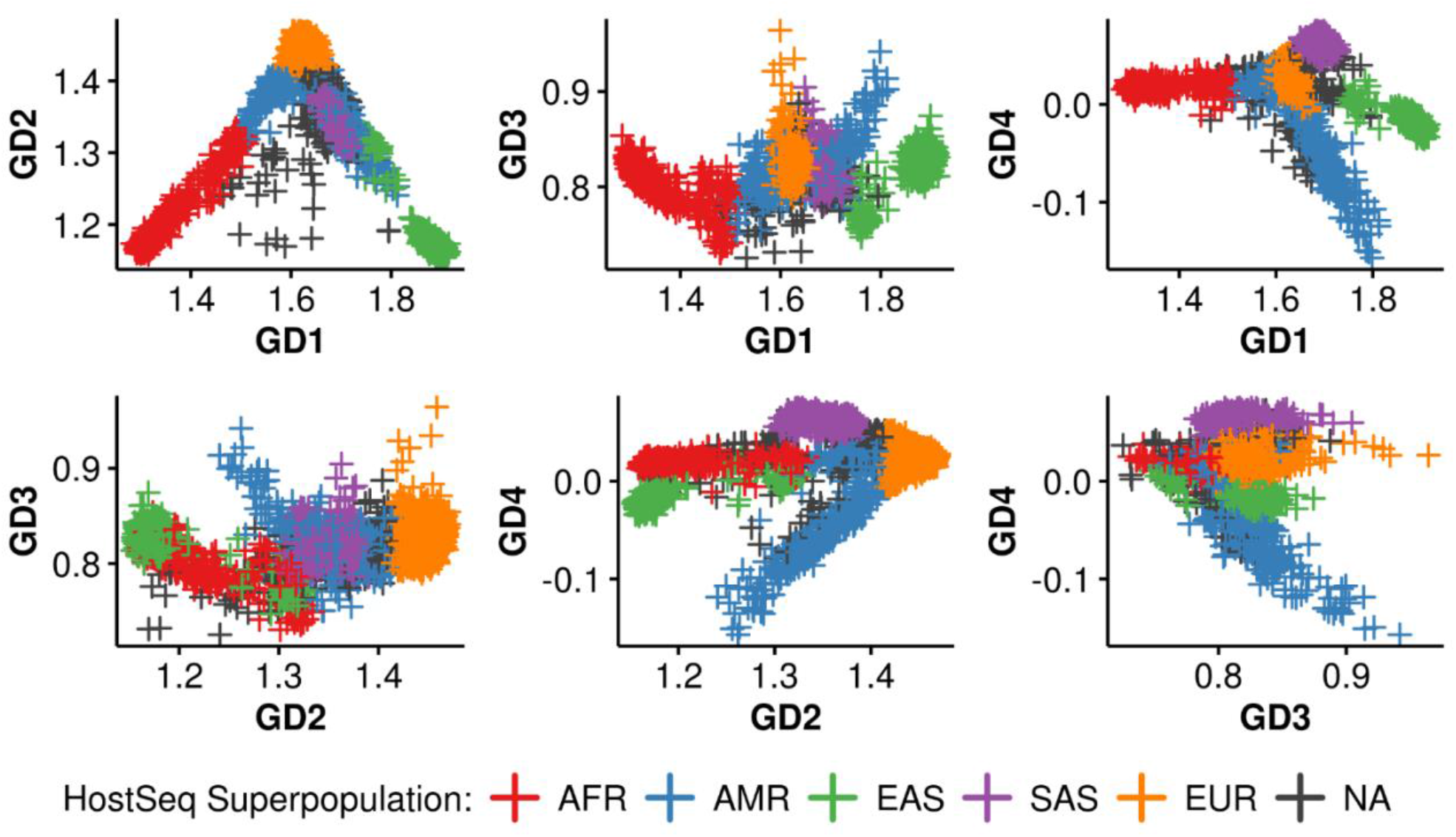
Genetic distances score of HostSeq genomes. The four genetic distances (GD1-4) scores from GRAF-pop (see Methods) represent distance of each genome from several reference populations and are used to predict ancestry. Barycentric coordinates of GD1 and GD2 are used to predict admixture proportion of African, East Asian and European ancestries.

## Footnotes

1 Aspects of graphics acquired from Wikimedia Commons.

2 https://ftp.1000genomes.ebi.ac.uk/vol1/ftp/technical/reference/GRCh38_reference_genome/

